# Rapid Genome Sequencing Compared to a Gene Panel in Critically Ill Infants with a Suspected Genetic Disorder: An Economic Evaluation

**DOI:** 10.1101/2024.10.18.24315740

**Authors:** Tara A. Lavelle, Jill L. Maron, Stephen F. Kingsmore, Ching-Hsuan Lin, Yingying Zhu, Benjamin Sweigart, Dallas Reed, Bruce D. Gelb, Jerry Vockley, Jonathan M. Davis

**Affiliations:** Center for the Evaluation of Risk in Health, Institute for Clinical Research and Health Policy Studies, Tufts Medical Center, 800 Washington St., #063, Boston, Massachusetts, 02111, USA.; Department of Medicine, Tufts University School of Medicine, Boston, MA; Women and Infants Hospital of Rhode Island, 101 Dudley St, Providence, Rhode Island 02905; Rady Children’s Institute for Genomic Medicine, 7910 Frost St Suite #300, San Diego, California, 92123; Tufts Clinical and Translational Science Institute, Tufts University, and Institute for Clinical Research and Health Policy Studies, Tufts Medical Center, 800 Washington Street, #63, Boston, Massachusetts 02111; Department of Pediatrics and Department of Obstetrics and Gynecology, 800 Washington St North Building, Tufts University School of Medicine, Boston, Massachusetts 02111; Department of Medicine, Tufts Medical Center, Boston, MA; Mindich Child Health and Development Institute and Departments of Pediatrics and Genetics and Genomic Sciences, Icahn School of Medicine at Mount Sinai, 1470 Madison Avenue Eighth Floor New York, New York 10029; University of Pittsburgh Medical Center Children’s Hospital, University of Pittsburgh School of Medicine, 4401 Penn Ave, Pittsburgh, Pennsylvania 15224; Department of Pediatrics, Tufts Medical Center, Boston, Massachusetts

## Abstract

**Introduction:** Rapid genome sequencing (rGS) provides high diagnostic yield for critically ill infants with suspected genetic disorders, but has high upfront costs and insufficient insurance coverage. Assessing the downstream costs and health outcomes associated with rGS is important for guiding coverage decisions. This study compares 1-year healthcare costs and quality-adjusted life years (QALYs) for: 1) early rGS (within 7 days of admission) for all infants, and 2) early targeted neonatal gene sequencing (NewbornDx) for all infants, followed by later rGS (after 7 days) for undiagnosed infants.

**Study Design:** The Genomic Medicine for Ill Neonates and Infants (GEMINI) study was a multicenter, prospective study that enrolled 400 hospitalized infants under one year of age with suspected genetic disorders. All participants underwent both rGS and NewbornDx. Using GEMINI data and 2023 Medicare rates, we developed a decision tree to compare total costs and QALYs over a 1-year period for the two testing strategies.

**Results:** The diagnostic yield and upfront testing costs were higher for rGS (49%; $12,297) than NewbornDx (27%; $2,449; p<0.05). As neither early testing nor diagnosis significantly affected QALYs, we conducted a cost-minimization analysis, focusing solely on cost differences between strategies. Over one year, early rGS was estimated to save $158,592 per patient (95% CI: $63,701-$253,292) compared to early NewbornDx with later rGS if necessary.

**Conclusions:** Early rGS results in substantial healthcare cost savings, highlighting the need to expand reimbursement to improve access early in a hospitalization for critically ill infants.

## Introduction

Genetic disorders are a major cause of infant mortality in the United States.^1^ For critically ill infants with suspected genetic disorders, traditional genetic tests are often ordered serially, have low diagnostic yields, and slow turnaround times, which can delay informed care management. In the U.S., rapid genome sequencing (rGS) can generally provide results within seven days and offers higher diagnostic yields, but higher upfront costs have limited its widespread use.^2, 3^ Previous U.S. economic evaluations of rGS in critically ill infants that included upfront and downstream costs have demonstrated that rGS is cost-effective or cost saving when used as a first-line diagnostic approach.^2, 4^ However, these studies have relied on data from restricted geographic areas, limited follow-up duration, or decision analytic models developed without empirical data.^2, 4^

The Genomic Medicine for Ill Neonates and Infants (GEMINI) study was the first multi-state U.S. prospective study of rGS in hospitalized, critically ill infants (age < 1 year) with suspected genetic disorders (ClinicalTrials.gov Identifier: <u>NCT03890679</u>).^5^ The study has been described in detail elsewhere.^5^ In summary, from June 2019 to November 2021, 400 enrolled participants underwent simultaneous testing with rGS (Rady Laboratory) and NewbornDx (Athena Diagnostics), a commercial targeted neonatal gene-sequencing panel that can identify variants across 1,722 genes linked to infant-onset disorders. The GEMINI study compared the tests’ time to result, diagnostic yield, and clinical utility of these two tests. Although enrollment depended on the proband, trio testing was preferred. Participants were followed from enrollment until 1 year corrected gestational age (CGA). Although NewbornDx offered faster median time to return of results (4.2 versus 6.1 days for rGS), rGS had a significantly higher diagnostic yield (49% versus 27%).^2, 3^

The GEMINI study also aimed to conduct the first evidence-based U.S. economic evaluation comparing rGS to NewbornDx in a diverse cohort of critically ill hospitalized infants with suspected genetic disorders. As rGS and NewbornDx were performed concurrently in GEMINI, patient-level data from the study was used to conduct a model-based economic evaluation comparing two hypothetical testing strategies: 1) early initiation of rGS for all hospitalized infants, and 2) early initiation of NewbornDx for all hospitalized infants followed by later rGS for undiagnosed infants. The goal of the study was to compare costs and quality adjusted life years (QALYs) between the two strategies over the one-year study period from the U.S. healthcare perspective.

## Methods

### Overview

This study included several stages: data collection during the GEMINI study, data analysis, and incorporation of the analyzed data into a decision tree, a type of decision analytic model. The study received approval from the Johns Hopkins Single Institutional Review Board (IRB; JHUSIRB00000007) and all participating site IRBs. The submission to the IRB included an economic evaluation study protocol in addition to the clinical study protocol. The economic evaluation study protocol is not publicly available. Informed consent was obtained from the parents of all participants.

### Data collection

Study site coordinators collected baseline data from families, including the infant’s gestational age, birth weight, enrollment age, sex, parents’ self-described race and ethnicity, and residential ZIP code to match to median household income.^6^ Caregivers reported follow-up healthcare utilization and health-related quality of life (HRQOL) data until the infant reached 1 year CGA. After discharge from the infant’s enrolling hospitalization, we contacted parents/caregivers monthly with questionnaires about their infant’s prior month healthcare utilization including hospitalizations, physician visits, laboratory tests, imaging, procedures, home services, allied health professional visits, and medications. Parents/caregivers also rated their infant’s HRQOL on a visual analog scale (VAS) from 0 (“worst health”) to 100 (“best health”) and their own HRQOL using the 12-item Short-Form Health Survey (SF-12) at discharge and every three months.^7^ When parents/caregivers did not complete the questionnaire electronically and were unreachable by phone, coordinators used the infant’s medical records to complete the resource utilization portion of the survey and left the HRQOL portion blank. For each parent/caregiver-reported hospitalization at the enrolling hospital, coordinators obtained de-identified hospital billing charges and Current Procedural Terminology (CPT)/Healthcare Common Procedure Coding System (HCPCS) codes for inpatient services and uploaded them into REDCap.^8, 9^

### Estimating Resource Utilization and Costs

All reported healthcare utilization and hospital charge data were converted into costs. For outpatient care, we assigned CPT/HCPCS codes and used the 2023 Centers for Medicare & Medicaid Services (CMS) Medicare Physician and Clinical Laboratory Fee Schedules to assign reimbursement rates to these codes as proxies for costs.^10, 11^ Although Medicare does not cover most pediatric patients, Medicare reimbursement rates closely mirror actual costs for many services. We reviewed all outpatient medications and excluded their costs from the analysis because they were relatively inexpensive generics which had minimal annual costs. When estimating the costs and length of stay of the enrolling hospitalization, we included any additional costs and length of stay related to transfers that occurred directly from the enrolling hospitalization to another acute-care hospital for acute or convalescent care. We converted inpatient hospital billing charges to costs using each enrolling hospital’s Medicare cost-to-charge ratio. Costs were adjusted to 2023 U.S. dollars using the U.S. medical care consumer price index.^12^ Due to the study’s short time horizon, we did not discount costs or health utility values.

### Estimating Health Utility

A validated algorithm was used to convert SF-12 results into Short Form 6- Dimensions (SF-6D) utility values, which are preference-based HRQOL values ranging from 0 (dead) to 1 (perfect health). We divided VAS scores by 100 to estimate utility scores and assigned a utility of 0 to deceased infants. Based on caregiver grief studies, we assigned a utility value of 0.64 to bereaved parents/caregivers following an infant’s death.^13^ We calculated accrued QALYs for infants and parents/caregivers from enrollment through 1 year CGA.^14^ QALYs weigh the duration of time spent in a health state by the utility value associated with that state. We calculated QALYs by weighting utility scores measured at each survey time point by the duration of each interval before and after data collection and summing across all intervals.

### Addressing Missing Data

The analysis used multiple imputation by chained equations to impute missing observations for healthcare resource use, non-enrolling hospitalization costs, utility values, and baseline demographic variables. The R package procedure generated thirty imputed datasets.^15, 16^ **Supplemental Methods 1** provides the detailed methods.

### Descriptive Statistics

We stratified our cohort based on the timing of their rGS and NewbornDx testing following hospital admission: 1) early testing (≤ 7 days after admission) and 2) later testing (> 7 days after admission). This 7-day cutoff was chosen reflected both the median time to testing in our cohort and clinical agreement that testing within the first week is critical for informing timely management in infants with suspected genetic disorders. Baseline characteristics and study outcomes were reported for the entire cohort and for each test group. For categorical data, we reported frequencies. For continuous data, we provided means and standard deviations (SD) or medians with interquartile ranges (IQR). To compare baseline characteristics between groups, the analysis used a t-test for normally distributed continuous variables, the Kruskal-Wallis test for skewed continuous variables, and the Fisher’s exact and chi-squared tests for categorical data (2-sided, 5% significance level).

### Identifying Factors Influencing Utilization, Costs, and Utility

*Healthcare utilization and costs*: We created three generalized linear models with log link functions to examine the influence of early versus later testing and diagnosis on enrolling hospital costs (model 1), enrolling hospital length of stay (model 2), and total costs over the study period (enrollment to 1 year CGA) (model 3). The models adjusted for baseline clinical and demographic characteristics including birth weight, gestational age, race, ethnicity, household income, and insurance. A sensitivity analysis additionally adjusted for death with and without the withdrawal of care to account for potential confounding from reduced hospitalization and study days. A second sensitivity analysis additionally adjusted for urgent testing, which may be an indicator for greater severity of illness. Urgent testing was performed for participants who required mechanical ventilation, had severe neurological injury, were hemodynamically unstable, or when requested by the site principal investigator.^5^ We trimmed total costs to the 97.5^th^percentile to limit the influence of outliers based on Cook’s distance.^17^ We calculated uncertainty intervals using the Multiple-imputation Boot method.^18^

*Utility*: A two-part model (logit followed by linear regression) examined the impact of early versus later testing and diagnosis on QALYs accrued by infants over the study period. We chose a two-part model due to the substantial number of infants in the study who had 0 QALYs due to their early death. A logit regression estimated the probability of accruing non-zero QALYs, the linear regression identified factors associated with QALYs gained among infants with non-zero QALYs. We investigated how early versus later testing and diagnosis influenced caregiver QALYs accrued over the study period using a linear regression model. The models adjusted for death without the withdrawal of care and all other covariates specified in the cost model. Statistical analyses were performed using STATA version 17 and R 4.3.0. (R Core Team, 2023).^19, 20^

### Economic Evaluation

The model-based economic evaluation compared costs across two hypothetical strategies:1) early rGS, and 2) early NewbornDx followed by later rGS for undiagnosed infants (early NewbornDx/later rGS). These strategies were selected to address ongoing questions about the optimal testing sequence for children with suspected genetic disorders and whether less expensive tests should be used prior to rGS. We used a decision tree (**Figure 1**) due to the short time horizon for the study and obtained data inputs from several sources (**Table 1**).

**Figure 1.**
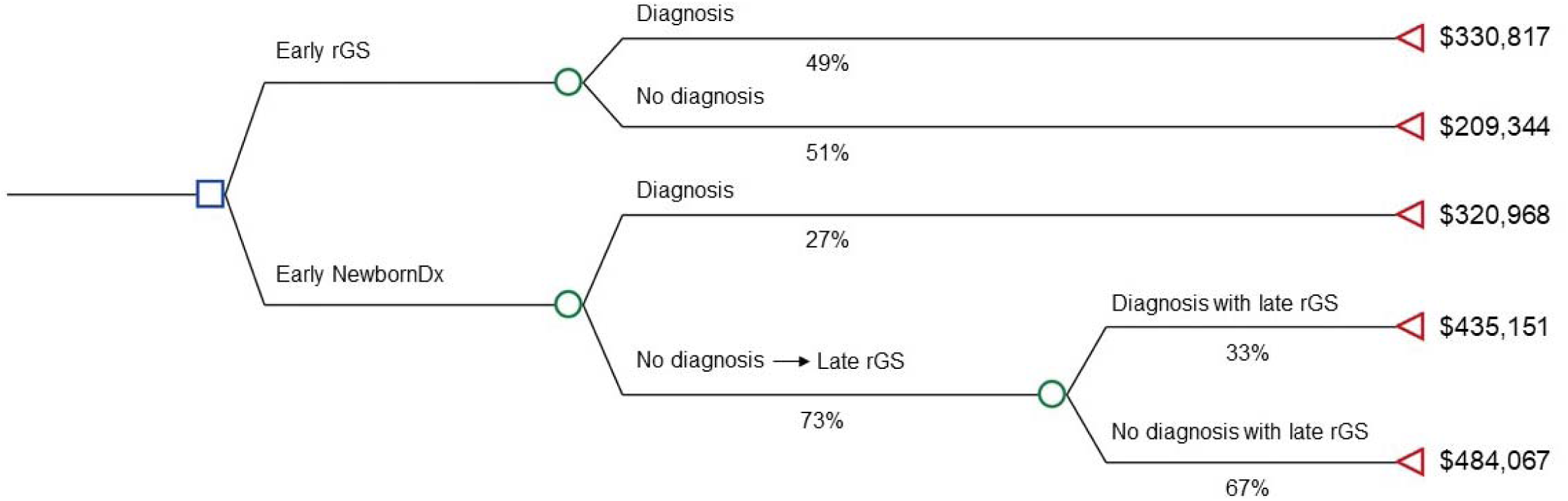
Decision analytic model comparing two hypothetical testing strategies for critically ill hospitalized infants with suspected genetic conditions

**Table 1.** Decision Analytic Model Inputs and Ranges Used in Probabilistic and Deterministic Sensitivity Analyses.

| Decision analytic model input | Mean<sup>a</sup> | Distributions used in probabilistic sensitivity analysis | 95% CI of distributions used in probabilistic sensitivity analysis | Ranges used in deterministic sensitivity analyses |
| --- | --- | --- | --- | --- |
| **Diagnostic rates** |  |  |  |  |
| Early rapid genome sequencing (rGS) | 0.49<sup>b</sup> | Binomial | (0.44, 0.54) | - |
| Early NewbornDx | 0.27 | Binomial | (0.23, 0.32) | - |
| Late rGS, following NewbornDx for undiagnosed infants | 0.33 | Binomial | (0.28, 0.39) | - |
| **Costs of rGS and NewbornDx<sup>a</sup>** |  |  |  |  |
| rGS | \$12,297.40 | | - | \$1,000 to \$12,297 |
| NewbornDx | \$2,448.56 | | - | \$100 to \$2,449 |
| **Other healthcare costs during the study period** |  |  |  |  |
| Early testing with a diagnosis, and no late testing | \$318,520 | Sampling distribution from GEMINI data | (\$220,108, \$429,139) | - |
| Early testing without a diagnosis, and no late testing | \$197,046 | Sampling distribution from GEMINI data | (\$138,211, \$263,891) | - |
| Late testing with a diagnosis (following early testing without a diagnosis) | \$420,405 | Sampling distribution from GEMINI data | (\$285,019, \$572,058) | - |
| Late testing without a diagnosis (following early testing without a diagnosis) | \$469,321 | Sampling distribution from GEMINI data | (\$358,768, \$587,167) | - |

<sup>a</sup> All model inputs were derived from GEMINI study data, except for: (1) rGS cost, which was derived using a weighted average of 2023 Medicare reimbursement rates for the CPT codes 0094U (proband) and 81426 (duo and trio) based on the rates of proband/duo/trio utilization in GEMINI, and (2) NewbornDx, was based on the 2023 Medicare reimbursement rate for the CPT code 81443.
<sup>b</sup> In the model, we changed the rGS diagnostic status of the one participant with Prader-Willi syndrome from non-diagnostic to diagnostic. We made his adjustment because the current diagnostic capabilities of rGS now can identify mutations associated with Prader-Willi syndrome. As such, the diagnostic rate for rGS changed from 48.75% to 49.00%.

*Diagnostic rates:* We obtained diagnostic rates from GEMINI study variant result classifications.^5^ Infants were classified as having a diagnosis if they had a diplotype that fit the pattern of inheritance of a disorder that comprised a variant or variants classified as pathogenic, likely pathogenic, or of unknown significance (VUS) only if associated with genes that fit the infant’s phenotype. All NewbornDx diagnoses were confirmed through rGS, except for nine infants where NewbornDx identified a diagnosis that rGS did not. We used this partial overlap to estimate the diagnostic rate for later rGS following a negative early NewbornDx result in the decision model.

*Costs of rGS and NewbornDx:* We used CPT codes and Medicare reimbursements rates to estimate the costs of NewbornDx and rGS, accounting for whether rGS was performed as a proband, duo, or trio.

*Other healthcare costs during the study period:* The decision analytic model includes four unique pathways across both strategies (**Figure 1**): 1) early testing with a diagnosis (no later testing), 2) early testing without a diagnosis (no later testing), 3) later testing (following early testing) with a diagnosis, and 4) later testing (following early testing) without a diagnosis. We used multivariable regression models to predict total healthcare costs for these four pathways, excluding the costs of rGS and NewbornDx.

We included all inputs in the decision model and calculated each strategy’s expected costs using R4.3.0 (R Core Team, 2023).^19^ To account for various sources of uncertainty, we performed a probabilistic sensitivity analysis (PSA). We used binomial distributions to characterize the uncertainty around the proportion of infants receiving a diagnosis from GS and/or NewbornDx and used bootstrapping with replacement to generate sampling distributions for total healthcare costs across the four model pathways. The PSA ran 10,000 iterations to construct 95% uncertainty intervals around our mean expected values. The PSA held rGS and NewbornDx costs constant and we used one- and two-way deterministic sensitivity analyses to estimate their contribution to uncertainty. In two additional scenario analyses we: 1) assumed that VUS results were non-diagnostic; 2) did not trim total costs during the study period.

## Results

### Sample Characteristics

**Table 2** presents baseline demographic and clinical characteristics of infants and families, comparing those tested early versus later. The median infant age at enrollment was 18.0 days (IQR:8.0-66.2) with 58% being male and 57% covered by public insurance. Early and later testers differed in terms of a number of characteristics, including their median age at enrollment [8.0 days (IQR: 4.0-65.0) versus 32.0 (IQR: 13.0-68.5); p<0.01], mean gestational age [37.2 weeks (SD:3.1) versus 36.0 (SD:4.0); p<0.01], and birth weight [2809 grams (SD: 817) versus 2596 (SD: 989), p=0.02].

**Table 2.**
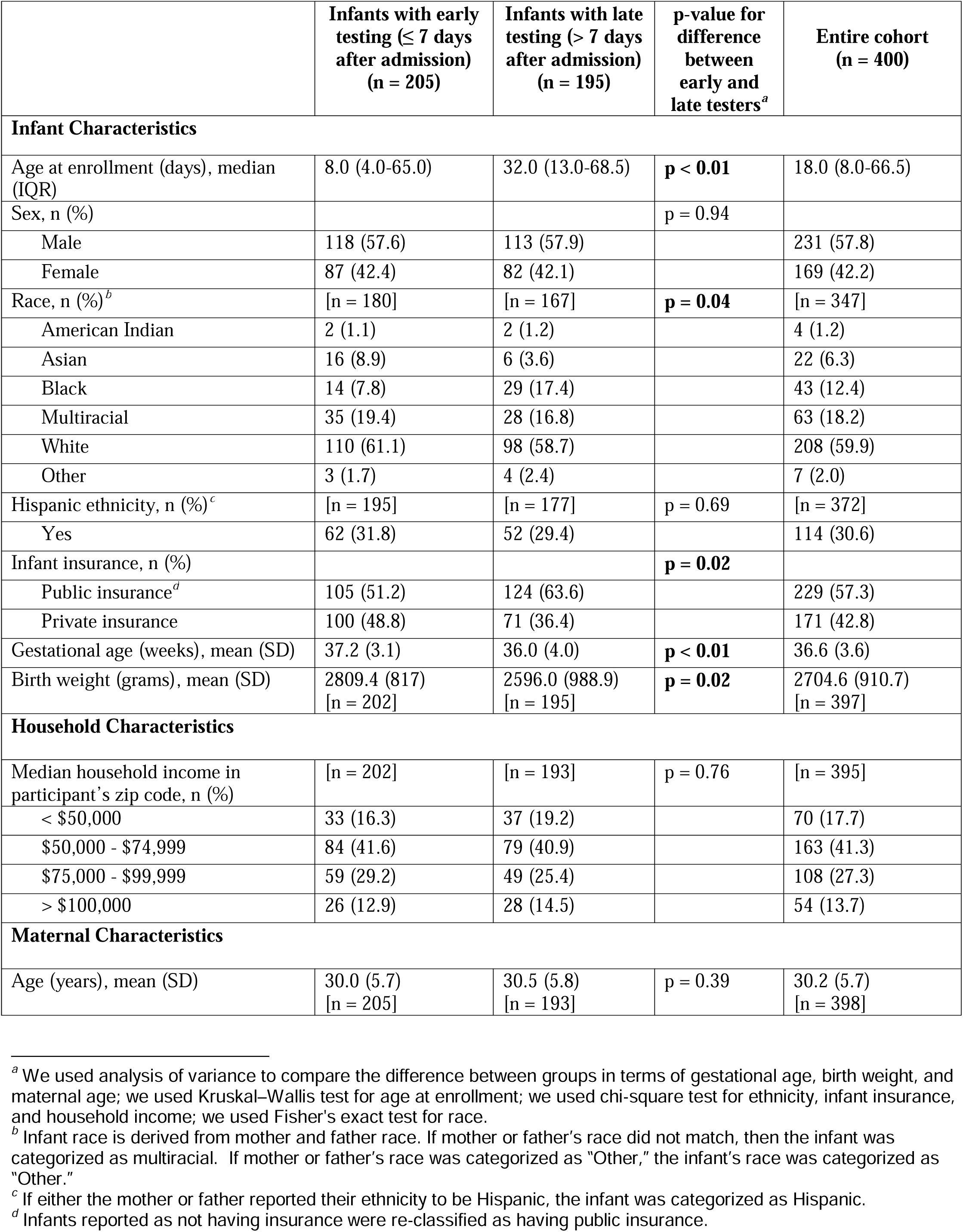
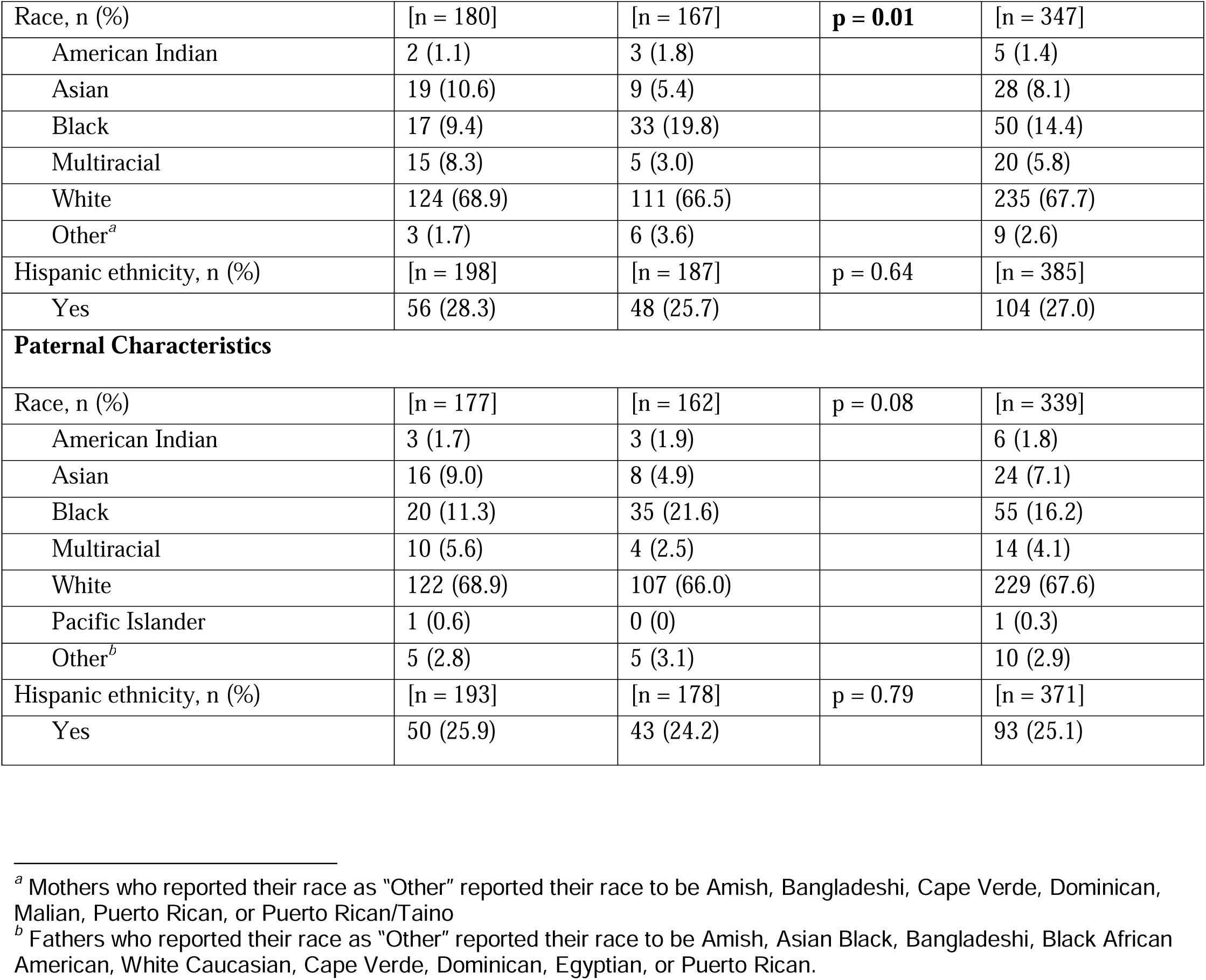
Infant and Family Baseline Characteristics.

### Survey response rates

A parent/caregiver or study coordinator completed 98% of healthcare utilization surveys.

Forty-four percent of parents/caregivers completed all child and parent HRQOL surveys, with 93% completing at least one survey. **Supplemental Table 1** displays characteristics of infants and families with and without missing HRQOL data.

### Unadjusted Resource Utilization and Costs During the Enrolling Hospitalization

Most infants (78%) received trio rGS, 18% infant-mother duo, and 4% proband. Fifty-one percent of infants underwent early testing. The mean enrolling hospitalization cost was $295,835 (SD: $25,770) per patient, with a median cost of $115,401 (IQR: $109,225-$123,038). The mean enrolling hospitalization length of stay was 60.8 days (SD: 74.3) with a median of 33.0 days (IQR: 14.0-79.0). Additional resource utilization is shown in **Table 3** and Supplemental Table 2.

**Table 3.** Unadjusted Study Outcomes.

|  | **Infants with early testing**<br>**(≤ 7 days after admission)**<br>(n = 205) | **Infants with late**<br>**testing (> 7 days after**<br>**admission)**<br>(n = 195) | **Entire cohort**<br>(n = 400) |
| --- | --- | --- | --- |
| **Diagnostic rates<sup>a</sup>** |  |  |  |
| Rapid genome sequencing (rGS) | 0.55 | 0.43 | 0.49 |
| NewbornDx | 0.32 | 0.22 | 0.27 |
| rGS and NewbornDx (used in combination) | 0.57 | 0.46 | 0.51 |
| **Enrolling hospitalization testing and length of stay** |  |  |  |
| Type of rGS, n (%) |  |  |  |
| Proband | 5 (2.4) | 11 (5.6) | 16 (4.0) |
| Duo | 30 (14.6) | 42 (21.5) | 72 (18.0) |
| Trio | 170 (82.9) | 142 (72.8) | 312 (78.0) |
| Total length of stay for enrolling hospitalization (days) |  |  |  |
| Mean (SD) | 36.4 (48.8) | 86.5 (87.0) | 60.8 (74.3) |
| Median (IQR) | 20.0 (9.0-40.0) | 58.0 (25.5-119.5) | 33.0 (14.0-79.0) |
| NICU length of stay for enrolling hospitalization (days)<sup>b</sup> |  |  |  |
| Mean (SD) | 31.2 (43.3) [n = 205] | 71.6 (68.8) [n = 194] | 50.9 (60.6) [n = 399] |
| Median (IQR) | 17.0 (5.0-36.0) [n = 205] | 52.5 (21.0-103.0) [n = 194] | 29.0 (10.0-69.0) [n = 399] |
| Discharge location<sup>c</sup>, n (%) |  |  |  |
| Home | 166 (81.0) | 147 (75.4) | 313 (78.3) |
| Another inpatient hospital (Transfer) | 6 (2.9) | 11 (5.6) | 17 (4.3) |
| Long term care facility | 0 (0.0) | 3 (1.5) | 3 (0.8) |
| Inpatient rehabilitation facility | 2 (1.0) | 5 (2.6) | 7 (1.8) |
| Infant died prior to discharge | 30 (14.6) | 25 (12.8) | 55 (13.8) |
| Hospitalized until 1 year CGA<sup>d</sup> | 1 (0.5) | 4 (2.1) | 5 (1.3) |
| **Follow-up resource utilization: discharge from enrolling hospitalization to 1 year CGA<sup>e</sup>** |  |  |  |
| Duration of follow-up period (days) | [n = 205] | [n = 195] | [n = 400] |
| Mean (SD) | 260.5 (147.5) [n = 205] | 280.4 (129.3) [n = 195] | 270.2 (139.1) |
| Median (IQR) | 336.0 (119.0-368.0) | 339.0 (195.0-357.0) | 339.0 (153.0-363.0) |
| Among infants who completed the study | [n = 154] | [n = 158] | [n = 312] |
| Mean (SD) | 317.6 (108.0) | 319.6 (97.4) | 318.6 (102.6) |

<sup>a</sup> The diagnosis rates include variants categorized as pathogenic, likely pathogenic, or variants of unknown significance
<sup>b</sup> Includes any additional days that may have resulted from transfers to acute-care hospitals
<sup>c</sup> From the enrolling hospital
<sup>d</sup> Corrected gestational age

|  |  |  |  |
| --- | --- | --- | --- |
| Median (IQR) | 358.0 (264.8-380.0) | 349.0 (301.2-370.8) | 353.0 (290.5-375.0) |
| Among infants who were lost to follow up | [n = 9] | [n = 4] | [n = 13] |
| Mean (SD) | 254.9 (126.2) | 222.0 (124.2) | 244.8 (121.4) |
| Median (IQR) | 274.0 (196.0-341.0) | 231.5 (156.5-297.0) | 274.0 (186.0-341.0) |
| Among infants who died during the study | [n = 42] | [n = 33] | [n = 75] |
| Mean (SD) | 52.4 (71.1) | 99.9 (108.4) | 73.3 (91.9) |
| Median (IQR) | 27.5 (10.0-66.2) | 66.0 (22.0-138.0) | 35.0 (12.0-96.0) |
| Subsequent hospitalizations<sup>a</sup> | [n = 96] | [n = 96] | [n = 192] |
| Mean (SD) | 3.0 (2.6) | 2.8 (2.2) | 2.9 (2.4) |
| Median (IQR) | 2.0 (1.0-4.0) | 2.0 (1.0-4.0) | 2.0 (1.0-4.0) |
| Emergency room visits, mean (SD) | 1.8 (0.2) | 1.6 (0.2) | 1.71 (0.1) |
| Primary care visits, mean (SD) | 7.5 (0.3) | 7.6 (0.4) | 7.6 (0.2) |
| Specialist outpatient visits, mean (SD) | 10.3 (0.6) | 12.6 (0.9) | 11.4 (0.6) |
| Cardiologist | 1.2 (0.2) | 1.8 (0.4) | 1.5 (0.2) |
| Gastrointestinal | 0.9 (0.1) | 1.5 (0.2) | 1.2 (0.1) |
| Geneticist | 0.7 (0.1) | 0.6 (0.1) | 0.6 (0.1) |
| Neurologist | 1.3 (0.2) | 0.8 (0.1) | 1.1 (0.1) |
| Pulmonologist | 0.5 (0.1) | 1.0 (0.1) | 0.8 (0.1) |
| Other, mean (SD)<sup>b</sup> | 5.6 (0.5) | 6.8 (0.6) | 6.2 (0.4) |
| Complex care clinic visits, mean (SD) | 0.3 (0.1) | 0.3 (0.1) | 0.3 (0.04) |
| Outpatient labs, tests, or procedures, mean (SD) |  |  |  |
| Blood work | 4.7 (0.6) | 3.5 (0.4) | 4.1 (0.4) |
| Echocardiogram | 1.8 (0.2) | 1.6 (0.2) | 1.7 (0.1) |
| Genetic test | 0.9 (0.1) | 0.6 (0.1) | 0.8 (0.1) |
| MRI | 0.7 (0.1) | 0.6 (0.1) | 0.7 (0.1) |
| Outpatient surgery | 0.7 (0.1) | 0.6 (0.1) | 0.7 (0.1) |
| X-ray | 1.3 (0.2) | 1.1 (0.1) | 1.2 (0.1) |
| Other<sup>b</sup> | 11.3 (1.0) | 9.1 (0.8) | 10.2 (0.6) |
| Occupational, physical or speech therapy visits<sup>c</sup>, mean (SD) | 22.3 (1.6) | 24.6 (1.7) | 23.4 (1.2) |
| Home nursing visits, mean (SD) | 11.8 (2.7) | 14.7 (2.8) | 13.2 (2.0) |
| Home health aide visits, mean (SD) | 2.1 (0.4) | 2.2 (0.7) | 2.1 (0.4) |
| Long-term care (LTC) admissions, mean (SD) | 0.2 (0.2) | 0.6 (0.5) | 0.4 (0.2) |
| Inpatient rehabilitation admissions, mean (SD) | 1.4 (1.4) | 1.5 (1.1) | 1.4 (0.9) |
| **Total costs (2023 US\$)** | | | |

<sup>a</sup> Subsequent hospitalizations are any hospitalizations that occur after discharge from the enrolling hospitalization or after discharge from an acute-care hospital that the infant was transferred to from the enrolling hospital.
<sup>b</sup> See eTable 7 for details on “Other” resource utilization
<sup>c</sup> At home, in-person, or through telehealth

|  |  |  |  |
| --- | --- | --- | --- |
| **Enrolling hospitalization<sup>a</sup>** |  |  |  |
| Mean (SD) | \$194,072 (23,074) | \$402,990 (46,258) | \$295,835 (25,770) |
| Median (IQR) | \$77,934 (76,880-79,801) | \$191,117(171,326-199,516) | \$115,401 (109,225-123,038) |
| **Post-discharge follow-up** |  |  |  |
| Mean (SD) | \$70,282 (19,463) | \$116,463 (24,629) | \$92,912 (15,681) |
| Median (IQR) | \$34,577 (28,620-45,859) | \$49,147 (44,482-64,869) | \$45,350 (40,509-47,564) |
| **Total cost until infant's 1 year CGA<sup>b</sup>** |  |  |  |
| Mean (SD) | \$264,455 (29,615) | \$519,575 (54,436) | \$388,572 (31,271) |
| Median (IQR) | \$120,044 (106,951-130,492) | \$241,837 (217,103-262,902) | \$169,990 (156,216-179,758) |

<sup>a</sup> Includes any additional costs that may have resulted from direct transfers to acute-care hospitals
<sup>b</sup> Corrected gestational age

### Unadjusted Resource Utilization and Costs During Follow-up and Total Study Period

The mean follow-up period from enrolling hospitalization discharge to one year CGA was 270.2 days (SD: 139.1) with a median of 339.0 days (IQR: 153.0-363.0). During this period, infants had a mean of 2.9 subsequent hospitalizations (SD: 2.4), 7.6 primary care visits (SD: 0.2), and 11.4 specialty outpatient visits (SD: 0.6). The mean follow-up cost per patient was $92,912 (SD: $15,681) with a median of $45,350 (IQR: $40,509-$47,564). For the total study period, the mean total cost per patient was $388,572 (SD: $31,271) with a median of $169,990 (IQR: $156,216-$179,758). (**Table 3**).

### Adjusted Cost and Length of Stay Analyses

In multivariate regression analyses adjusting for baseline characteristics, early testing (irrespective of platform) was associated with significantly reduced costs and length of stay during the enrolling hospitalization versus later testing (mean per patient reductions of $189,719 and 54 days, respectively; p<0.01 for both estimates, **Tables 4 -5**). Over the entire study period, early testing was associated with a mean cost savings of $272,275 per patient (p<0.01) versus later testing **(Table 1)**. Diagnosis and its interaction with early testing did not significantly predict enrolling hospital costs, length of stay, or total costs (**Table 4**). Adjusting for infant death with and without withdrawal of care did not alter the findings (**Supplemental Table 3**). This indicates that after adjusting for other characteristics, enrolling hospitalization costs, length of stay, and total costs were not significantly different between infants who died and those who survived. Adjusting for urgent testing, which was more common within the early testing period (69% of early testers were urgent cases of versus 13% of later testers, p<0.05 for difference), also did not alter the findings (**Supplemental Table 4**).

**Table 4.** Multivariate Regression Models Predicting: 1) Enrolling Hospital Costs, 2) Enrolling Hospital Length of Stay (LOS), 3) Total Costs Over the Study Period.

| Independent variables | Outcome variables |  |  |  |  |  |
| --- | --- | --- | --- | --- | --- | --- |
|  | Model 1. Enrolling hospital cost |  | Model 2. Enrolling hospital LOS |  | Model 3. Total costs over study period |  |
|  | Exponentiated coefficient <sup>a</sup> | p-value | Exponentiated coefficient <sup>a</sup> | p-value | Exponentiated coefficient <sup>a</sup> | p-value |
| (Intercept) | \$467,918 | < **0.001** | 110 days | < **0.001** | \$575,191 | < **0.001** |
| Late testing (> 7 days after admission) (ref) |  |  |  |  |  |  |
| Early testing (≤ 7 days after admission) | 0.46 | **0.002** | 0.35 | < **0.001** | 0.43 | **0.002** |
| Infant did not receive a Diagnosis (ref) |  |  |  |  |  |  |
| Infant did receive a Diagnosis | 0.87 | 0.36 | 0.88 | 0.21 | 0.88 | 0.41 |
| (Interaction): Early testing * with Diagnosis | 1.56 | 0.15 | 1.64 | 0.07 | 1.77 | 0.07 |
| Normal or low birth weight (> 1500g) (ref) |  |  |  |  |  |  |
| Very low birth weight (≤ 1500g) | 1.75 | **0.02** | 1.54 | **0.01** | 1.41 | 0.18 |
| Gestational age at birth: > 32 wks (ref) |  |  |  |  |  |  |
| Gestational age at birth: ≤ 32 wks | 0.91 | 0.69 | 0.93 | 0.70 | 0.97 | 0.91 |
| Household income: < \$50,000 (ref) | | | | | | |
| \$50,000 - \$74,999 | 0.68 | **0.01** | 0.78 | **0.02** | 0.76 | 0.08 |
| \$75,000 - \$99,999 | 0.77 | 0.11 | 0.67 | **0.002** | 0.77 | 0.12 |
| ≥ \$100,000 | 0.76 | 0.23 | 0.68 | **0.03** | 0.68 | 0.14 |
| Infant race and ethnicity: White, non-Hispanic or Latino (ref) |  |  |  |  |  |  |
| Black, non-Hispanic or Latino | 1.15 | 0.42 | 1.09 | 0.46 | 1.40 | 0.06 |
| Hispanic or Latino | 0.95 | 0.74 | 0.93 | 0.51 | 1.04 | 0.81 |
| Other race, non-Hispanic or Latino | 0.63 | 0.11 | 0.92 | 0.62 | 0.75 | 0.30 |
| Insurance: Public (ref) |  |  |  |  |  |  |
| Private | 0.85 | 0.26 | 0.89 | 0.25 | 0.91 | 0.54 |

<sup>a</sup> Coefficients have been exponentiated to show the multiplicative change in the outcome variable for different categories of the predictor variable relative to the reference category. The intercept represents the baseline outcome variable and the reference category of all predictors.

**Table 5.** Predicted Enrolling Hospital Costs and Length of Stay across the Four Model Pathways.

|  | Enrolling hospital cost |  | Enrolling hospital length of stay (days) |  |
| --- | --- | --- | --- | --- |
|  | Mean | 95% uncertainty intervals | Mean | 95% uncertainty intervals |
| **Early testing with a diagnosis, and no late testing** | \$215,220 | (\$143,519, \$295,565) | 42 | (31, 55) |
| **Early testing without a diagnosis, and no late testing** | \$154,259 | (\$106,851, \$214,076) | 29 | (22, 35) |
| **Late testing with a diagnosis (following early testing without a diagnosis)** | \$304,098 | (\$217,283, \$405,325) | 73 | (60, 88) |
| **Late testing without a diagnosis (following early testing without a diagnosis)** | \$343,978 | (\$265,584, \$426,800) | 83 | (70, 97) |

Based on the multivariate regression model, the expected mean per patient costs over the total study period were: 1) early testing with a diagnosis: $318,520 (95% CI: $220,108-$429,139), 2) early testing without a diagnosis: $197,046 (95% CI: $138,211-$263,891), 3) later testing with a diagnosis: $420,405 (95% CI: $285,019-$572,058), and 4) later testing without a diagnosis: $469,321 (95% CI: $358,768-$587,167) (**Table 1**). As noted with the overlapping 95% confidence intervals, these results are consistent with the regression findings that cost differences were significant between early and later testing, but not between those who did or did not receive a diagnosis.

### Economic Evaluation

Because neither early testing nor diagnosis had a significant impact QALYs (**Table 6, Supplemental Table 5**), we conducted a cost-minimization analysis, focusing solely on cost differences between strategies rather than differences in health outcomes.

**Table 6.** Unadjusted Quality-Adjusted Life Years (QALYs) Accrued over the Study Period.

| **Sample** | **Infants with early testing (≤ 7 days after admission)**<br>**(n = 179)** | **Infants with late testing ($>$ 7 days after admission)**<br>**(n = 174)** | **Entire cohort included in QALY analysis<sup>a</sup>**<br>**(n = 353)** |
| --- | --- | --- | --- |
| **Infants** | 0.57 | 0.62 | 0.59 |
| **Caregivers** | 0.70 | 0.71 | 0.70 |

<sup>a</sup> Twenty-seven infants and twenty-seven caregivers were excluded from the analysis because their HRQOL values were missing across all time points

The expected total costs per patient over the study period are $269,566 (95% CI: $211,194-$332,326) for early rGS and $428,158 (95% CI: $358,729-$503,403) for early NewbornDx/later rGS. Early rGS is expected to save $158,592 (95% CI: $63,701-$253,292) in healthcare costs per patient over the study period compared to NewbornDx/later rGS (**Table 7, Supplemental Figures 1-3 for sampling distributions**).

**Table 7.** Expected Total Costs Over the Study Period From the Decision Model That Compared Two Hypothetical Testing Strategies for Critically Ill Hospitalized Infants With Suspected Genetic Conditions.

| **Strategy** | **Cost** | **95% uncertainty interval** |
| --- | --- | --- |
| Early rapid genome sequencing (rGS) ($\leq 7$ days after hospital admission) | \$269,566 | (\$211,194, \$332,326) |
| Early NewbornDx ($\leq 7$ days after hospital admission), followed by later ($> 7$ days following admission) rGS for undiagnosed infants | \$428,158 | (\$358,729, \$503,403) |
| **Difference** | -\$158,592 | (-\$253,292, -\$63,701) |

Sensitivity analyses showed that varying the testing costs minimally affected results (**Supplemental Figures 4-5**). When VUS results were considered non-diagnostic, early rGS saved an estimated $186,816 (95% CI: $11,945-$373,887) compared to early NewbornDx/later rGS over the study period (**Supplemental Table 6)**. Without cost trimming in our regression models, early rGS saved $185,221 (95% CI: $47,677-$321,086) compared to early NewbornDx/later rGS over the study period (**Supplemental Table 6**).

## Discussion

Our decision analytic model demonstrates that performing early rGS for critically ill infants with suspected genetic disorders in the U.S. could save over $150,000 in healthcare costs during an infant’s first year of life compared to early NewbornDx followed by rGS (if needed). The cost savings stemmed primarily from shorter length of stay if later rGS was avoided. These results are consistent with prior studies showing that rGS is most cost-effective when used early in a hospitalization as a first-line test.^2, 4^ Importantly, this study leverages empirical evidence from GEMINI’s 400 infants and offers insights into resource utilization spanning from the enrolling hospitalization through one year of follow-up, a timeframe pertinent for U.S. payer decision-making.

Currently, most state Medicaid programs (covering approximately 40% of U.S. children) and private insurers do not routinely reimburse hospitals for inpatient rGS.^21^ Our findings are timely as more payers are considering or are being required by new legislation to establish supplemental payments to hospitals for inpatient rGS.^2^ Separate reimbursement for rGS will enable all eligible patients to access rGS early in a hospitalization, maximizing its clinical and economic benefits while minimizing health disparities.

Our study highlights the economic value of ordering rGS early in a hospitalization, regardless of diagnostic outcome. This result aligns with GEMINI, which reported that physicians rated rGS very useful/useful in 87% of diagnosed and 65% of undiagnosed cases.^5^ One benefit of early rGS is it permits physicians to make timely and informed care management decisions based on the presence or absence of genetic diagnosis.

One study limitation is the lack of recorded health outcome data post-testing, aside from self-reported infant and caregiver HRQOL used to estimate accrued QALYs. While QALYs are standard in health economic evaluations, they may not adequately capture the benefit of rGS in infants with suspected genetic disorders. One goal of rGS is to identify genetic disorders for which therapeutic interventions may extend life and improve HRQOL. Another goal is to provide parents with prognostic information that may lead to the withdrawal of life-sustaining support, reducing QALYs. Given these limitations, we recommend that future economic evaluations of rGS also use objective health outcome measures post-testing. These measures are crucial for understanding the clinical implications of molecular diagnoses and assessing the economic impact of subsequent treatments and management strategies. Future studies should also extend follow-up periods to evaluate whether evolving phenotypic data can influence diagnostic yield and economic impact. Extended time horizons are also needed to capture long-term clinical outcomes, HRQOL, and the full economic value of genomic testing as its impact unfolds over time.

Another limitation of our study is that testing time was not randomized in GEMINI. To evaluate the independent association between predictor variables (timing of test initiation and diagnosis) and model outcomes (cost, length of stay, and QALYs), we used regression models that adjusted for all available baseline infant and household demographic and clinical characteristics. We also controlled for infant death and urgent testing in two separate sensitivity analyses, which demonstrated that both of these variables were not confounders in our adjusted analyses. However, unobserved or unrecorded data that may correlate with early testing, diagnosis, and outcomes could lead to potential confounding and bias in our estimates.

We also relied on parent-reported healthcare utilization or abstraction of this information from medical records, which may not fully reflect the true healthcare use for the infant. Finally, our model used empirical data from the GEMINI study that had broad entry criteria and variable enrollment rates across sites. Optimizing the use of rGS could further improve its diagnostic and economic value, especially as further therapeutic options are developed to treat rare genetic and metabolic disorders in neonates.

In GEMINI, some children with complex clinical presentations had “partial” diagnoses, both from rGS and NewbornDx, meaning the identified genetic variant explained only part of their clinical phenotype. For simplicity, we combined “partial” and “full” diagnoses into a single “diagnosis” category in the model. This simplification provides conservative cost estimates for early NewbornDx, as infants partially diagnosed by NewbornDx were classified as “diagnosed” and thus were assumed not to require later rGS. If, in practice, infants with partial diagnoses from NewbornDx still undergo later rGS, this would increase the overall costs of the early NewbornDx strategy, thereby further highlighting the cost-savings associated with early rGS.

The model also did not include comparisons to exome sequencing (ES) or other clinically available genetic tests besides NewbornDx. Previous studies have shown that GS has a higher diagnostic rate and is more cost-effective than ES or other genetic tests when it is used as a first-line test.^4, 22^

We conducted this analysis from the U.S. healthcare system perspective to inform payer coverage decisions for rGS. However, this approach does not capture broader societal costs and benefits associated with rGS, such as caregiver productivity, family out-of-pocket expenses for non-healthcare utilization, and personal utility, which is the utility associated with testing that is not health related. Personal utility may be influenced by the perceived benefits of receiving rGS results, such as improved coping, feeling a sense of control, and the ability to plan for the future.^23^ Conversely, personal utility may also be affected by concerns about rGS including potential discrimination and privacy risks.^24–29^ Although societal costs and benefits could affect the overall economic value of rGS, they were outside the scope of this study and should be considered in future economic evaluations.

In addition, this study was conducted at six academic medical centers, which may impact the generalizability of its findings to other settings. Future research should take place in a broader range of healthcare settings, including community hospitals and rural healthcare facilities, to inform whether the findings from this study are consistent across different environments.

Other important factors related to the appropriate utilization of rGS are also noteworthy, and economic value is only one of many considerations.^30^ Ethical, legal, and social implications also play a critical role in determining the appropriateness and impact of rGS.^23, 31^ For example, ethical concerns include issues of equity and access to early rGS. Legal factors include the implementation of laws to protect the privacy of genetic information. Social implications include the psychological impact of testing and diagnoses on families and children as they age. These factors, in combination with health economic data, should be considered to make informed, responsible decisions about the appropriate implementation of rGS in clinical practice.^30^

Critically ill infants with suspected genetic disorders who undergo early rGS have significantly lower costs compared to those who receive early NewbornDx, primarily due to the frequent reflex to rGS. This underscores the urgent need for payers to ensure timely and equitable access to rGS for all eligible infants, thereby improving access to this important diagnostic tool, maximizing clinical and economic benefits, and reducing health disparities.

## Data Availability

Data and models are available from the corresponding author upon request.

