## Supplemental for "Rapid Genome Sequencing Compared to a Gene Panel in Critically Ill Infants with a Suspected Genetic Disorder: An Economic Evaluation"

### Supplemental Online Content


**Supplemental Methods 1.** Description of the Multivariate Imputation by Chained Equations (MICE) Procedure

**Supplemental Table 1.** Sample Characteristics by Missing Health-Related Quality of Life (HRQOL) Survey Status and Inclusion or Exclusion in Quality-Adjusted Life Year (QALY) Analyses

**Supplemental Table 2.** Other Resource Utilization over the Entire Study Period

**Supplemental Table 3.** Sensitivity Multivariate Regression Model Results Including Variables Controlling for Infant Death With and Without Withdrawal of Care. Models Predict: 1) Enrolling Hospital Costs, 2) Enrolling Hospital Length of Stay (LOS), 3) Total Costs over the Study Period

**Supplemental Table 4.** Sensitivity Multivariate Regression Model Results Including Variable Controlling for Urgent Testing. Models Predict: 1) Enrolling Hospital Costs, 2) Enrolling Hospital Length of Stay (LOS), 3) Total Costs Over the Study Period

**Supplemental Table 5.** Regression Models for Infant and Caregiver Quality-Adjusted Life Years (QALYs)

**Supplemental Figure 1.** Probabilistic Sensitivity Analysis for the Total Cost over the Study Period for Early rGS Based on 10,000 Simulation Iterations of the Decision Analytic Model

**Supplemental Figure 2.** Probabilistic Sensitivity Analysis for the Total Cost over the Study Period for Early NewbornDx Based on 10,000 Simulation Iterations of the Decision Analytic Model

**Supplemental Figure 3.** Probabilistic Sensitivity Analysis for the Total Cost Saving over the Study Period for Early rGS Compared to Early NewbornDx Based on 10,000 Simulation Iterations of the Decision Analytic Model

**Supplemental Figure 4.** Deterministic Sensitivity Analysis: One-Way Sensitivity Analysis Examining How the Total Cost Savings Associated with rGS Change When the Costs of rGS and NewbornDx Are Varied from Low to High Values in the Decision Analytic Model

**Supplemental Figure 5.** Deterministic Sensitivity Analysis of the Decision Analytic Model: Two-Way Sensitivity Analysis Examining How the Total Cost Savings Associated with rGS Change When the Costs of rGS and NewbornDx Are Simultaneously Varied from Low to High Values

**Supplemental Table 6.** Additional Sensitivity Analyses of Model Inputs and Assumptions

**Supplemental Methods 1. Description of the Multivariate Imputation by Chained Equations (MICE) Procedure**

To address missing data, we used multivariate imputation by chained equations (MICE). Below, we outline the MICE setup, including the number of imputed datasets, the cycles performed per imputation, the variables used for predicting missing values, and bounds applied during imputation.
We conducted 10 cycles of iteration for each imputed dataset. We generated 30 datasets for healthcare resource utilization, hospitalization costs from non-enrolling hospitals, utility values, infant birth weight, ZIP code-matched median household income, and gestational age.

**Predictor variables included:**
- Gestational age
- Birth weight
- Diagnostic results
- Medication interventions (added, removed, or changed)
- Surgical interventions (added, removed, or changed)
- Dietary changes
- Care goals (redirected from comfort to cure)
- Any change in management or goals of care
- New tests ordered or previous tests canceled
- Screening for additional comorbidities (added or removed)
- New imaging sought or prior imaging canceled
- New specialty services sought or previous services no longer required
- Change in cure
- Mother's race and ethnicity
- Infant's insurance
- Median income based on birth ZIP code using American Community Survey (ACS) data

Additional variables were included for the imputation of hospitalization costs from non-enrolling hospitals and utility values:

**Hospitalization costs from non-enrolling hospitals:**
- Length of stay

**Health care utilization:**
- Total number of primary care visits over the study period
- Total number of specialist visits over the study period
- Total number of complex care visits over the study period
- Total number of hospitalization days over the study period

We specified ranges for imputing some missing values in healthcare resource utilization. For example, if the parent reported that their infant received a specific healthcare service, but did not indicate the quantity, we assigned an imputed value greater than 0 for the quantity. If the parent reported that their child visited the emergency room but did not indicate how many times, we imputed a value between 1 and the maximum expected number of emergency room visits from the survey.
Thirteen infants lost to follow-up were excluded from cost imputation. Utility values were not imputed for 27 individuals who were alive but missing data at all five time points.

**Supplemental Table 1. Sample Characteristics by Missing Health-Related Quality of Life (HRQOL) Survey Status and Inclusion or Exclusion in Quality-Adjusted Life Year (QALY) Analyses**

| | **EXCLUDED**<br><br>**Infants who did not die during study but had no HRQOL data**<br><br>**(n = 27)** | **INCLUDED**<br><br>**Infants who died during enrolling hospitalization**<br><br>**(n = 58)** | **INCLUDED**<br><br>**Infants with some HRQOL data reported (with the remaining data imputed)**<br><br>**(n = 198)** | **INCLUDED**<br><br>**Infants with all HRQOL data reported (no imputations)**<br><br>**(n = 117)** | **p-values: comparing infants excluded to infants included in the analysis** |
|---|---|---|---|---|---|
| Time of testing | | | | | 0.21 |
| Early testers, n (%) | 17 (63.0) | 32 (55.2) | 102 (51.5) | 54 (46.2) | |
| Late testers, n (%) | 10 (37.0) | 26 (44.8) | 96 (48.5) | 63 (53.8) | |
| Received a diagnosis | | | | | **0.021** |
| Yes, n (%) | 8 (29.6) | 36 (62.1) | 100 (49.5) | 60 (48.7) | |
| No, n (%) | 19 (70.4) | 22 (37.9) | 98 (49.5) | 57 (48.7) | |
| **Infant Characteristics** | | | | | |
| Gestational age (weeks), mean (SD) | 36.8 (3.8) | 35.8 (3.2) | 36.9 (3.6) | 36.5 (3.8) | 0.748 |
| Birth weight (gram), mean (SD) | 2706.2 (943.2) [n = 27] | 2399.9 (798.7) [n = 56] | 2787.6 (911) [n = 198] | 2709.9 (932.7) [n = 116] | 0.992 |
| Age at enrollment (days), median (IQR) | 41 (8-81) | 12 (4-37.8) | 18 (8-64) | 24 (8-85) | 0.197 |
| Sex, n (%) | | | | | 0.811 |
| Male | 15 (55.6) | 32 (55.2) | 118 (59.6) | 66 (56.4) | |
| Female | 12 (44.4) | 26 (44.8) | 80 (40.4) | 51 (43.6) | |
| Race, n (%)[a] | | | | | **0.009** |
| American Indian | 0 (0.0) | 1 (1.7) | 1 (0.5) | 2 (1.7) | |
| Asian | 2 (7.4) | 5 (8.6) | 10 (5.1) | 5 (4.3) | |
| Black | 2 (7.4) | 9 (15.5) | 22 (11.1) | 10 (8.5) | |
| Multiracial | 11 (40.7) | 8 (13.8) | 31 (15.7) | 13 (11.1) | |
| White | 7 (25.9) | 23 (39.7) | 106 (53.5) | 72 (61.5) | |
| Other | 5 (18.5) | 12 (20.7) | 28 (14.1) | 15 (12.8) | |
| Hispanic ethnicity, n (%)[b] | 10 (37.0) | 16 (27.6) | 58 (29.3) | 30 (25.6) | 0.309 |
| Infant insurance, n (%) | | | | | 0.534 |
| Public insurance [c] | 17 (63.0) | 35 (60.3) | 115 (58.1) | 62 (53.0) | |
| Private insurance | 10 (37.0) | 23 (39.7) | 83 (41.9) | 55 (47.0) | |
| **Household Characteristics** | | | | | |
| Median household income in participant's zip code, n (%) | [n=26] | [n=57] | [n=197] | [n=115] | 0.087 |

[a] Infant race is derived from mother and father race. If mother or father's race did not match, then the infant was categorized as multiracial. If mother or father's race was categorized as "Other," the infant's race was categorized as "Other."
[b] If either the mother or father reported their ethnicity to be Hispanic, the infant was categorized as Hispanic.
[c] Infants reported as not having insurance were re-classified as having public insurance.

| | | | | | |
|---|---|---|---|---|---|
| < $50,000 | 1 (3.8) | 16 (28.1) | 35 (17.8) | 18 (15.7) | |
| $50,000 - $74,999 | 14 (53.8) | 19 (33.3) | 83 (42.1) | 47 (40.9) | |
| $75,000 - $99,999 | 5 (19.2) | 14 (24.6) | 52 (26.4) | 37 (32.2) | |
| ≥ $100,000 | 6 (23.1) | 8 (14.0) | 27 (13.7) | 13 (11.3) | |
| **Maternal Characteristics** | | | | | |
| Age (years), mean (SD) | 31.5 (5.5) [n = 27] | 29.0 (5.4) [n = 57] | 30.0 (6.1) [n = 198] | 31 (5.1) [n = 116] | 0.241 |
| Race, n (%) | | | | | 0.142 |
| American Indian | 0 (0.0) | 1 (1.7) | 1 (0.5) | 3 (2.6) | |
| Asian | 3 (11.1) | 5 (8.6) | 14 (7.1) | 6 (5.1) | |
| Black | 4 (14.8) | 9 (15.5) | 26 (13.1) | 11 (9.4) | |
| Multiracial | 4 (13.8) | 3 (5.2) | 11 (5.6) | 2 (1.7) | |
| White | 11 (40.7) | 28 (48.3) | 117 (59.1) | 79 (67.5) | |
| Other[a] | 5 (18.5) | 12 (20.7) | 29 (14.6) | 16 (13.7) | |
| Hispanic ethnicity, n (%) | 9 (37.5) [n=24] | 14 (25.9) [n=54] | 53 (27.5) [n=193] | 28 (24.6) [n=114] | 0.059 |
| **Paternal Characteristics** | | | | | |
| Race, n (%) | | | | | 0.229 |
| American Indian | 0 (0.0) | 1 (1.7) | 3 (1.5) | 2 (1.7) | |
| Asian | 3 (11.1) | 5 (8.6) | 12 (6.1) | 5 (4.3) | |
| Black | 5 (18.5) | 10 (17.2) | 28 (14.1) | 12 (10.3) | |
| Multiracial | 3 (11.1) | 3 (5.2) | 5 (2.5) | 3 (2.6) | |
| White | 11 (40.7) | 24 (41.4) | 119 (60.1) | 75 (64.1) | |
| Other[b] | 5 (18.5) | 15 (25.9) | 31 (15.7) | 20 (17.1) | |
| Hispanic ethnicity, n (%) | 9 (34.6) [n=26] | 13 (25) [n=52] | 47 (25.5) [n=184] | 24 (22.0) [n=109] | 0.378 |

[a] Mothers who reported their race as “Other” or “Unknown,” were categorized as “Other.”
[b] Fathers who report their race to be “Other,” “Unknown,” or “Pacific Islanders” were categorized as “Other”.

**Supplemental Table 2. Other Resource Utilization over the Entire Study Period**

| Resource | Infants with early testing (≤ 7days after admission) (n = 205) | Infants with late testing (> 7 days after admission) (n = 195) | Entire cohort (n = 400) |
|---|---|---|---|
| **Specialist outpatient visits, mean (SD)** | | | |
| Cardiologist | 1.2 (0.2) | 1.8 (0.4) | 1.5 (0.2) |
| Neurologist | 1.3 (0.2) | 0.8 (0.1) | 1.1 (0.1) |
| Developmental and Behavioral Pediatrician | 0.2 (0.1) | 0.5 (0.2) | 0.3 (0.1) |
| Gastrointestinal | 0.9 (0.1) | 1.5 (0.2) | 1.2 (0.1) |
| Hematologist | 0.7 (0.2) | 0.5 (0.2) | 0.6 (0.1) |
| Surgeon | 0.4 (0.1) | 0.6 (0.1) | 0.5 (0.1) |
| Ear-Nose-Throat Doctor | 0.6 (0.1) | 0.6 (0.1) | 0.6 (0.1) |
| Geneticist | 0.7 (0.1) | 0.6 (0.1) | 0.6 (0.1) |
| Pulmonologist | 0.5 (0.1) | 1.0 (0.1) | 0.8 (0.1) |
| Audiologist | 0.5 (0.1) | 0.3 (0.1) | 0.4 (0.1) |
| Endocrinologist | 0.3 (0.1) | 0.4 (0.1) | 0.3 (0.0) |
| Neonatologist | 0.4 (0.1) | 0.3 (0.1) | 0.3 (0.0) |
| Nephrologist | 0.2 (0.1) | 0.7 (0.2) | 0.5 (0.1) |
| Neurosurgeon | 0.3 (0.1) | 0.4 (0.1) | 0.4 (0.1) |
| Ophthalmologist | 0.6 (0.1) | 0.6 (0.1) | 0.6 (0.1) |
| Orthopedic Doctor/Surgeon | 0.5 (0.1) | 0.7 (0.2) | 0.6 (0.1) |
| Urologist | 0.2 (0.1) | 0.3 (0.1) | 0.3 (0.0) |
| General Surgeon | 0.3 (0.1) | 0.4 (0.1) | 0.4 (0.0) |
| Dietitian and Nutritionist | 0.1 (0.0) | 0.2 (0.0) | 0.2 (0.0) |
| Dermatologist | 0.1 (0.0) | 0.1 (0.0) | 0.1 (0.0) |
| Immunologist | 0.0 (0.0) | 0.1 (0.0) | 0.1 (0.0) |
| Anesthesiologist | 0.0 (0.0) | 0.0 (0.0) | 0.0 (0.0) |
| Chiropractor | 0.0 (0.0) | 0.0 (0.0) | 0.0 (0.0) |
| Colon Rectal Surgeon | 0.0 (0.0) | 0.0 (0.0) | 0.0 (0.0) |
| Infectious Disease Doctor | 0.0 (0.0) | 0.1 (0.0) | 0.0 (0.0) |
| Plastic Surgeon | 0.0 (0.0) | 0.1 (0.0) | 0.1 (0.0) |
| Physiologist | 0.0 (0.0) | 0.0 (0.0) | 0.0 (0.0) |
| Radiologist | 0.0 (0.0) | 0.0 (0.0) | 0.0 (0.0) |
| Rehabilitation Doctor | 0.0 (0.0) | 0.0 (0.0) | 0.0 (0.0) |
| Rheumatologist | 0.0 (0.0) | 0.0 (0.0) | 0.0 (0.0) |
| Palliative care | 0.0 (0.0) | 0.0 (0.0) | 0.0 (0.0) |
| **Complex care clinic visits, mean (SD)** | | | |
| Craniofacial Clinic | 0.0 (0.0) | 0.0 (0.0) | 0.0 (0.0) |
| Complex Congenital Heart Clinic | 0.1 (0.1) | 0.1 (0.0) | 0.1 (0.0) |
| Genetics Clinic | 0.0 (0.0) | 0.0 (0.0) | 0.0 (0.0) |
| Cerebral Palsy Clinic | 0.0 (0.0) | 0.0 (0.0) | 0.0 (0.0) |
| Spina Bifida Clinic | 0.0 (0.0) | 0.0 (0.0) | 0.0 (0.0) |
| Unknown type | 0.1 (0.0) | 0.1 (0.0) | 0.1 (0.0) |

| | | | |
|---|---|---|---|
| Aerodigestive clinic | 0.1 (0.0) | 0.1 (0.0) | 0.1 (0.0) |
| NICU Follow-up Clinic | 0.0 (0.0) | 0.0 (0.0) | 0.0 (0.0) |
| High Risk Follow-Up Clinic | 0.0 (0.0) | 0.0 (0.0) | 0.0 (0.0) |
| Muscle Disease Clinic | 0.0 (0.0) | 0.0 (0.0) | 0.0 (0.0) |
| **Outpatient labs, tests, or procedures, mean (SD)** | | | |
| Blood work | 4.7 (0.6) | 3.5 (0.4) | 4.1 (0.4) |
| Echo (echocardiogram) | 1.8 (0.2) | 1.6 (0.2) | 1.7 (0.1) |
| X-ray | 1.3 (0.2) | 1.1 (0.1) | 1.2 (0.1) |
| MRI | 0.7 (0.1) | 0.6 (0.1) | 0.7 (0.1) |
| Outpatient surgery | 0.7 (0.1) | 0.6 (0.1) | 0.7 (0.1) |
| Genetic test | 0.9 (0.1) | 0.6 (0.1) | 0.8 (0.1) |
| Fluoroscopy | 0.0 (0.0) | 0.0 (0.0) | 0.0 (0.0) |
| Fiberoptic Endoscopic Evaluation of Swallow/ FEES | 0.1 (0.0) | 0.1 (0.0) | 0.1 (0.0) |
| Electroencephalogram (EEG) | 0.2 (0.0) | 0.1 (0.0) | 0.1 (0.0) |
| Electrocardiogram (ECG or EKG) | 0.4 (0.1) | 0.3 (0.1) | 0.4 (0.1) |
| CT | 0.1 (0.0) | 0.1 (0.0) | 0.1 (0.0) |
| Respiratory Syncytial Virus Vaccine | 0.1 (0.0) | 0.0 (0.0) | 0.1 (0.0) |
| Culture | 0.0 (0.0) | 0.0 (0.0) | 0.0 (0.0) |
| Audiogram/Routine Hearing Test/ Hearing test/ Diagnostic Hearing Screen | 0.0 (0.0) | 0.0 (0.0) | 0.0 (0.0) |
| Urinalysis | 0.1 (0.0) | 0.1 (0.0) | 0.1 (0.0) |
| Sleep Study | 0.1 (0.0) | 0.1 (0.0) | 0.1 (0.0) |
| Auditory Brainstem Response (ABR) test | 0.0 (0.0) | 0.0 (0.0) | 0.0 (0.0) |
| Gastrostomy Tube/ Nasogastric Tube/ Feeding Tube Replacement | 0.0 (0.0) | 0.0 (0.0) | 0.0 (0.0) |
| Biopsy | 0.0 (0.0) | 0.0 (0.0) | 0.0 (0.0) |
| Bronchoscope | 0.0 (0.0) | 0.0 (0.0) | 0.0 (0.0) |
| Orthopedic Cast | 0.0 (0.0) | 0.0 (0.0) | 0.0 (0.0) |
| Holter monitor | 0.1 (0.0) | 0.0 (0.0) | 0.0 (0.0) |

**Supplemental Table 3. Sensitivity Multivariate Regression Model Results Including Variables Controlling for Infant Death With and Without Withdrawal of Care. Models Predict: 1) Enrolling Hospital Costs, 2) Enrolling Hospital Length of Stay (LOS), 3) Total Costs over the Study Period**

| **Independent variables** | **Outcome variables** | | | | | |
|---|---|---|---|---|---|---|
| | **Model 1. Enrolling hospital cost** | | **Model 2. Enrolling hospital LOS** | | **Model 3. Total study cost** | |
| | Exponentiated coefficient [a] | p-value | Exponentiated coefficient [a] | p-value | Exponentiated coefficient [a] | p-value |
| (Intercept) | $445,660 | < **0.001** | 103 days | < **0.001** | $544,726 | < **0.001** |
| Late testing (> 7 days after admission) (ref) | | | | | | |
| Early testing (≤ 7 days after admission) | 0.47 | < **0.01** | 0.35 | < **0.001** | 0.44 | < **0.01** |
| Infant did not receive a Diagnosis (ref) | | | | | | |
| Infant did receive a Diagnosis | 0.89 | 0.45 | 0.91 | 0.34 | 0.99 | 0.92 |
| (Interaction): Early testing * with Diagnosis | 1.62 | 0.12 | 1.67 | 0.06 | 1.76 | 0.08 |
| Normal or low birth weight (> 1500g) (ref) | | | | | | |
| Very low birth weight (≤ 1500g) | 1.75 | **0.02** | 1.55 | < **0.01** | 1.30 | 0.27 |
| Gestational age at birth: > 32 wks (ref) | | | | | | |
| Gestational age at birth ≤ 32 wks | 0.93 | 0.75 | 0.94 | 0.74 | 1.03 | 0.92 |
| Household income: < $50,000 (ref) | | | | | | |
| $50,000 - $74,999 | 0.70 | **0.02** | 0.80 | **0.04** | 0.74 | **0.04** |
| $75,000 - $99,999 | 0.79 | 0.16 | 0.68 | < **0.01** | 0.74 | 0.07 |
| ≥ $100,000 | 0.78 | 0.28 | 0.70 | **0.04** | 0.64 | 0.08 |
| Infant race and ethnicity: White, non-Hispanic or Latino (ref) | | | | | | |
| Black, non-Hispanic or Latino | 1.14 | 0.45 | 1.08 | 0.55 | 1.37 | < **0.05** |
| Hispanic or Latino | 0.97 | 0.86 | 0.95 | 0.66 | 1.08 | 0.60 |
| Other race, non-Hispanic or Latino | 0.61 | 0.09 | 0.92 | 0.60 | 0.70 | 0.19 |
| Insurance: Public (ref) | | | | | | |
| Private | 0.84 | 0.23 | 0.89 | 0.25 | 0.98 | 0.91 |

[a] Coefficients have been exponentiated to show the multiplicative change in the outcome variable for different categories of the predictor variable relative to the reference category. The exponentiated intercept represents the baseline outcome variable and the reference category of all predictors.

| | | | | | | |
|---|---|---|---|---|---|---|
| No death during enrolling hospitalization (Models 1 and 2) or study period (Model 3) (ref) | | | | | | |
| Death due withdrawal of care | 1.13 | 0.50 | 1.20 | 0.12 | 1.06 | 0.69 |
| Death due to reasons other than withdrawal of care | 0.58 | 0.46 | 0.46 | 0.36 | 0.42 | 0.34 |

**Supplemental Table 4. Sensitivity Multivariate Regression Model Results Including Variable Controlling for Urgent Testing. Models Predict: 1) Enrolling Hospital Costs, 2) Enrolling Hospital Length of Stay (LOS), 3) Total Costs Over the Study Period**

| Independent variables | Outcome variables | | | | | |
|---|---|---|---|---|---|---|
| | Model 1. Enrolling hospital cost | | Model 2. Enrolling hospital LOS | | Model 3. Total costs over study period | |
| | Exponentiated coefficient [a] | p-value | Exponentiated coefficient [a] | p-value | Exponentiated coefficient [a] | p-value |
| (Intercept) | $469,983 | < 0.001 | 110 days | < 0.001 | $573,716 | < 0.001 |
| Late testing (> 7 days after admission) (ref) | | | | | | |
| Early testing (≤ 7 days after admission) | 0.46 | 0.002 | 0.34 | < 0.001 | 0.45 | 0.004 |
| Infant did not receive a Diagnosis (ref) | | | | | | |
| Infant did receive a Diagnosis | 0.87 | 0.37 | 0.88 | 0.19 | 0.97 | 0.84 |
| (Interaction): Early testing * with Diagnosis | 1.57 | 0.15 | 1.63 | 0.07 | 1.71 | 0.10 |
| Normal or low birth weight (> 1500g) (ref) | | | | | | |
| Very low birth weight (≤ 1500g) | 1.75 | 0.02 | 1.55 | 0.01 | 1.32 | 0.23 |
| Gestational age at birth: > 32 wks (ref) | | | | | | |
| Gestational age at birth: ≤ 32 wks | 0.92 | 0.72 | 0.93 | 0.67 | 1.02 | 0.92 |
| Household income: < $50,000 (ref) | | | | | | |
| $50,000 - $74,999 | 0.68 | 0.01 | 0.77 | 0.02 | 0.73 | 0.03 |
| $75,000 - $99,999 | 0.76 | 0.10 | 0.67 | 0.003 | 0.69 | 0.03 |

[a] Coefficients have been exponentiated to show the multiplicative change in the outcome variable for different categories of the predictor variable relative to the reference category. The intercept represents the baseline outcome variable and the reference category of all predictors.

| | | | | | | |
|---|---|---|---|---|---|---|
| ≥ $100,000 | 0.76 | 0.23 | 0.68 | 0.03 | 0.62 | 0.0531 |
| Infant race and ethnicity: White, non-Hispanic or Latino (ref) | | | | | | |
| Black, non-Hispanic or Latino | 1.15 | 0.43 | 1.09 | 0.48 | 1.37 | 0.049 |
| Hispanic or Latino | 0.96 | 0.79 | 0.91 | 0.45 | 1.08 | 0.63 |
| Other race, non-Hispanic or Latino | 0.63 | 0.11 | 0.92 | 0.62 | 0.73 | 0.23 |
| Insurance: Public (ref) | | | | | | |
| Private | 0.86 | 0.29 | 0.88 | 0.21 | 1.02 | 0.89 |
| Urgent testing: No (ref) | | | | | | |
| Yes | 0.94 | 0.72 | 1.09 | 0.50 | 0.78 | 0.17 |

**Supplemental Table 5. Regression Models for Infant and Caregiver Quality-Adjusted Life Years (QALYs)**

| Independent variables | Outcome variables | | | | | |
|---|---|---|---|---|---|---|
| | Infant QALY, odds of having QALY > 0 | | Infant QALY, linear regression for infants with QALY > 0 | | Caregiver QALY, linear regression | |
| | Odds Ratio | p-value | Linear Coefficient | p-value | Linear Coefficient | p-value |
| Intercepts | 72.9 | **< 0.001** | 0.79 | **< 0.001** | 0.79 | **< 0.001** |
| Late testing (>7 days after hospital admission) (ref) | | | | | | |
| Early testing (≤7 days after hospital admission) | 1.2 | 0.840 | 0.01 | 0.804 | -0.01 | 0.648 |
| Infant did not receive a diagnosis (ref) | | | | | | |
| Infant did receive a diagnosis | 1.5 | 0.582 | -0.01 | 0.706 | -0.005 | 0.799 |
| (Interaction): Early testing * with diagnosis | 0.3 | 0.211 | -0.04 | 0.358 | 0.001 | 0.972 |
| Low or normal birth weight (>1500g) (ref) | | | | | | |
| Very low birth weight (≤ 1500g) | 5.6 | 0.054 | 0.07 | 0.299 | 0.04 | 0.215 |
| Gestational age at birth: >32 wks (ref) | | | | | | |
| Gestational age at birth: ≤ 32 wks | 0.1 | **0.017** | 0.10 | 0.129 | 0.09 | **0.005** |
| Household income: <$50,000 (ref) | | | | | | |
| $50,000 - $74,999 | 1.5 | 0.495 | -0.03 | 0.390 | 0.01 | 0.789 |
| $75,000 - $99,999 | 1.5 | 0.516 | 0.02 | 0.573 | 0.01 | 0.493 |
| ≥$100,000 | 0.5 | 0.384 | -0.06 | 0.208 | 0.01 | 0.588 |
| Infant race: White, non-Hispanic/Latino (ref) | | | | | | |
| Black, non-Hispanic/Latino | 0.4 | 0.186 | -0.04 | 0.394 | 0.03 | 0.118 |
| Hispanic or Latino | 0.4 | 0.109 | 0.03 | 0.255 | 0.01 | 0.368 |
| Other | 1.0 | 0.973 | 0.04 | 0.288 | 0.04 | 0.055 |
| Infant's insurance: Public (ref) | | | | | | |
| Private | 0.6 | 0.294 | -0.03 | 0.214 | -0.01 | 0.355 |
| Infant alive, or infant death w/ withdrawal of care during index hospitalization (ref) | | | | | | |
| Infant death, no withdrawal of care during index hospitalization | 0.6 | **< 0.001** | -0.43 | **< 0.001** | -0.13 | **< 0.001** |

Overall, 16.4% (N=58) of infants in this study (N=353) had zero infant QALYs because of death during the initial hospitalization. We, therefore, conducted two-part models to separately examine factors associated with the odds of having non-zero infant QALYs and with actual changes in infant QALYs among those with non-zero QALYs.

**Supplemental Figure 1. Probabilistic Sensitivity Analysis for the Total Cost over the Study Period for Early rGS Based on 10,000 Simulation Iterations of the Decision Analytic Model**

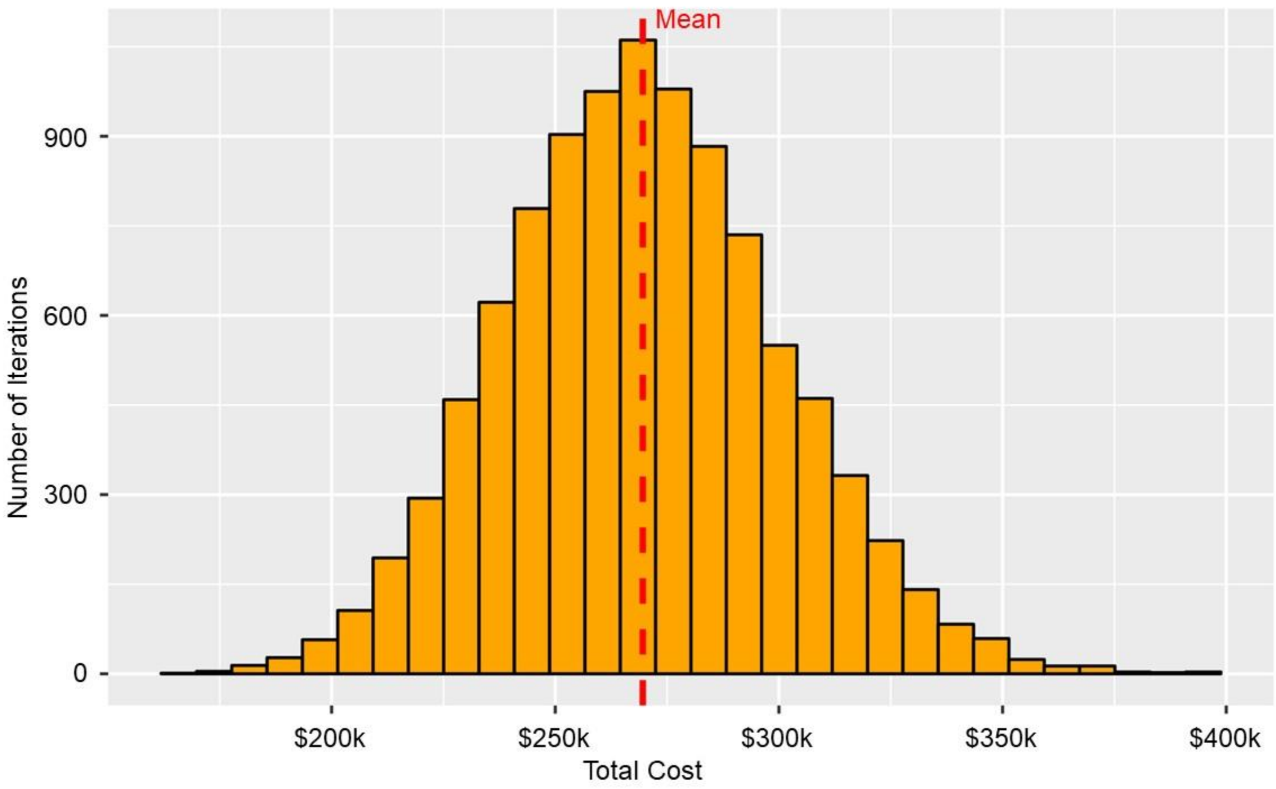

**Supplemental Figure 2. Probabilistic Sensitivity Analysis for the Total Cost over the Study Period for Early NewbornDx Based on 10,000 Simulation Iterations of the Decision Analytic Model**

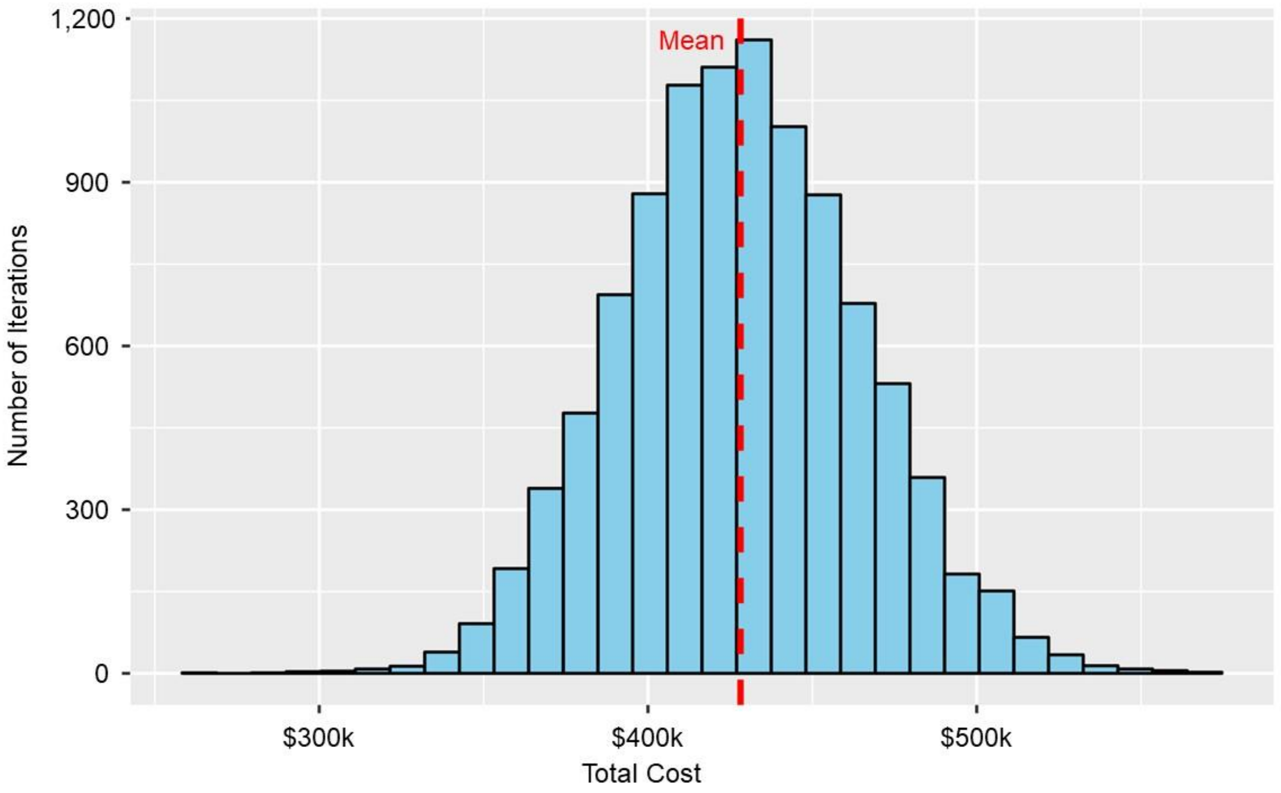

**Supplemental Figure 3. Probabilistic Sensitivity Analysis for the Total Cost Saving over the Study Period for Early rGS Compared to Early NewbornDx Based on 10,000 Simulation Iterations of the Decision Analytic Model**

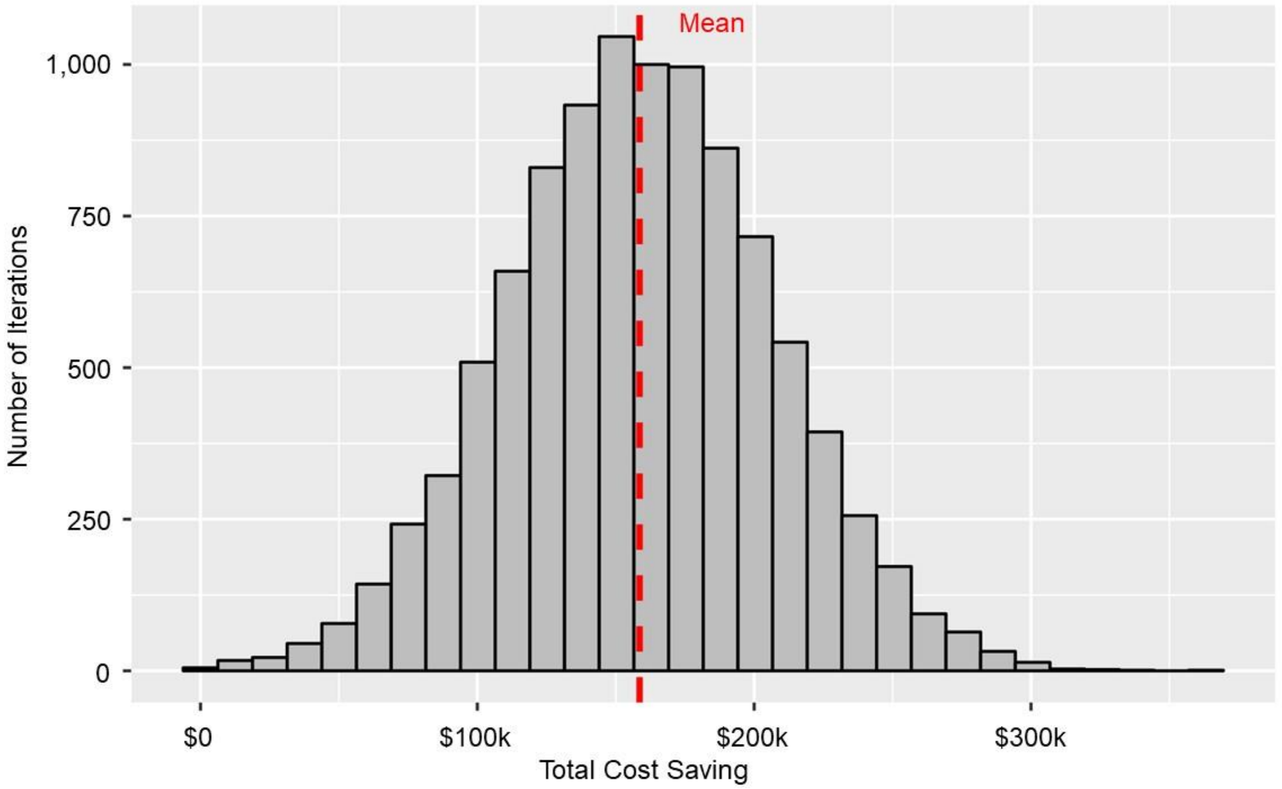

**Supplemental Figure 4. Deterministic Sensitivity Analysis: One-Way Sensitivity Analysis Examining How the Total Cost Savings Associated with rGS Change When the Costs of rGS and NewbornDx Are Varied from Low to High Values in the Decision Analytic Model**

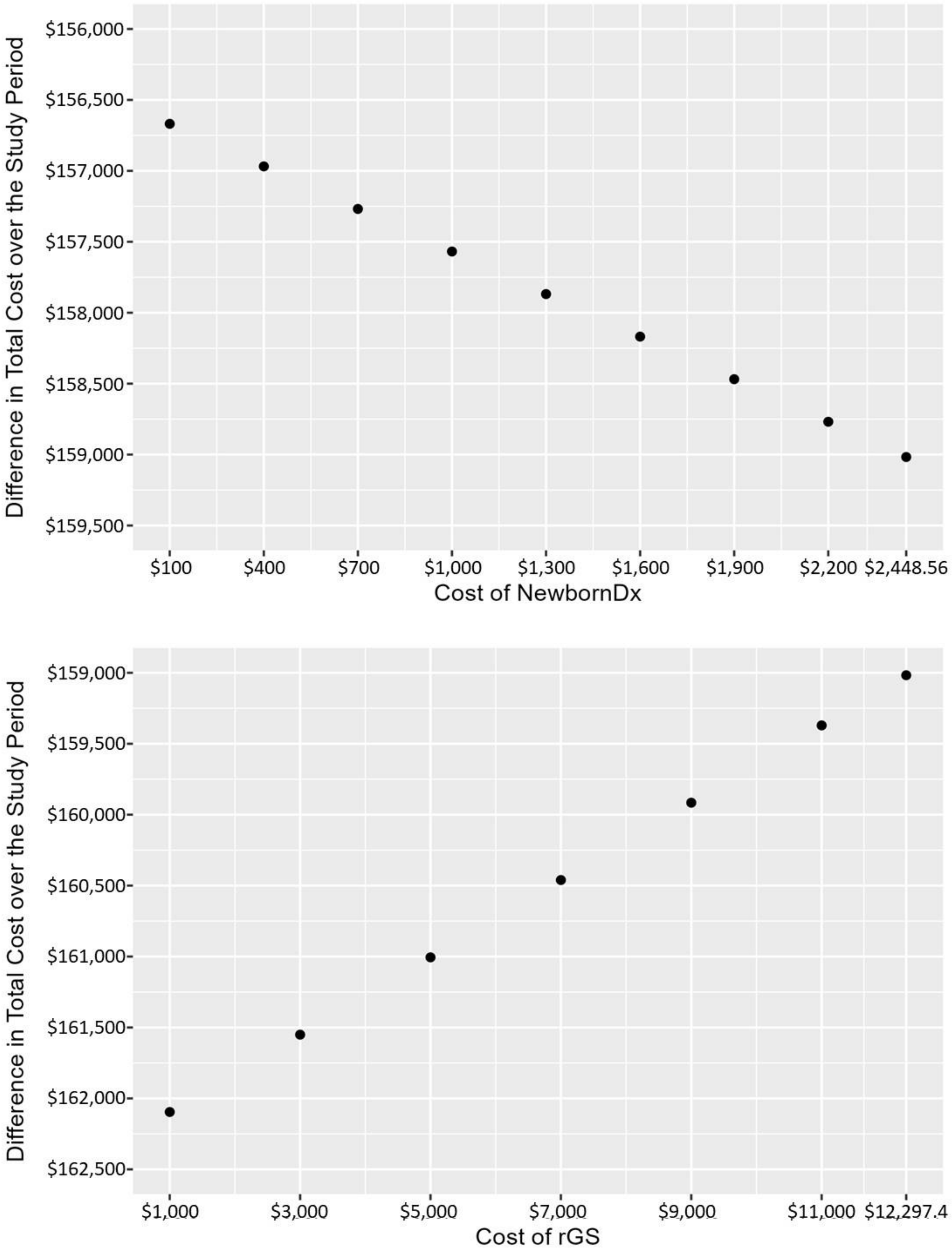

**Supplemental Figure 5. Deterministic Sensitivity Analysis of the Decision Analytic Model: Two-Way Sensitivity Analysis Examining How the Total Cost Savings Associated with rGS Change When the Costs of rGS and NewbornDx Are Simultaneously Varied from Low to High Values**

| | | cost of NewbornDx | | | | | | | | |
|---|---|---|---|---|---|---|---|---|---|---|
| | | $100 | $400 | $700 | $1,000 | $1,300 | $1,600 | $1,900 | $2,200 | $2,449 |
| cost of rGS | $1,000 | 159747 | 160047 | 160347 | 160647 | 160947 | 161247 | 161547 | 161847 | 162096 |
| | $3,000 | 159202 | 159502 | 159802 | 160102 | 160402 | 160702 | 161002 | 161302 | 161551 |
| | $5,000 | 158657 | 158957 | 159257 | 159557 | 159857 | 160157 | 160457 | 160757 | 161006 |
| | $7,000 | 158112 | 158412 | 158712 | 159012 | 159312 | 159612 | 159912 | 160212 | 160461 |
| | $9,000 | 157567 | 157867 | 158167 | 158467 | 158767 | 159067 | 159367 | 159667 | 159916 |
| | $11,000 | 157022 | 157322 | 157622 | 157922 | 158222 | 158522 | 158822 | 159122 | 159371 |
| | $12,297 | 156669 | 156969 | 157269 | 157569 | 157869 | 158169 | 158469 | 158769 | 159017 |

Colors correspond to the cost difference between early rGS and early NewbornDx

: Early rGS saves more than $160,000 compared to early NewbornDx.

: Early rGS saves between $160,000 and $159,000 compared to early NewbornDx.

: Early rGS saves between $159,000 and $158,000 compared to early NewbornDx.

: Early rGS saves less than $158,000 compared to early NewbornDx.

**Supplemental Table 6. Additional Sensitivity Analyses of Model Inputs and Assumptions**

| Model strategy | Base Case, mean (95% uncertainty intervals) | Assuming the VUS[a] results are not considered diagnostic[b], mean (95% uncertainty intervals) | No trimming of cost data, mean (95% uncertainty intervals) |
|---|---|---|---|
| Early rGS (≤ 7 days after hospital admission) | $269,566 ($211,194, $332,326) | $266,038 ($175,942, $362,822) | $260,543 ($182,406, $342,009) |
| Early NewbornDx (≤ 7 days after hospital admission), followed by late (> 7 days following admission) rGS for undiagnosed infants | $428,158 ($358,729, $503,403) | $452,855 ($175,942, $362,822) | $445,764 ($332,969, $555,654) |
| **Difference** | -$158,592 (-$63,701, -$253,292) | -$186,816 (-$11,945, -$373,887) | -$185,221 (-$47,677, -$321,086) |

[a] Variant of unknown significance

[b] If VUS results are not considered diagnostic, then early rGS would have a diagnostic rate of 37%, early Newborn diagnostic rate would be 20%, and late rGS (if no NewbornDx diagnosis) would have a diagnostic rate of 23%
